# Machine Learning to Investigate Life-Course Social Determinants of Loneliness among Older Adults in the US, England, Israel, and 27 European Countries during the Pandemic

**DOI:** 10.1101/2025.10.07.25337355

**Authors:** Xu Zong, Pekka Martikainen, Yuxuan Wang, Ling Liu, Karri Silventoinen

## Abstract

**Background:** Loneliness in later life is common and shaped by social determinants, with the COVID-19 pandemic and regional contexts further influencing disparities. Yet the role of life-course social determinants in predicting loneliness during the pandemic across countries remains unclear.

**Methods:** We used data from the Health and Retirement Study, the English Longitudinal Study of Ageing, and the Survey of Health, Ageing and Retirement in Europe, including 47,016 participants aged 65+ (mean age: 75). Surveys were conducted before and during the pandemic (April–August 2020). Six machine learning (ML) algorithms incorporating 18 predictors spanning childhood, pre-pandemic, and pandemic periods were applied to predict loneliness. Model optimization and interpretability employed cross-validation, hyperparameter tuning, and SHAP values.

**Findings:** CatBoost showed the best performance, with AUC scores of 0.718 in Europe and Israel, 0.693 in England, and 0.632 in the US. Key predictors common across regions included being single, female, parental education, and COVID-19-related adversity. Regional differences emerged: pre-pandemic income and financial support during the pandemic were stronger predictors in the US; receipt of public pension and parental occupation at age 10 were more influential in England; and parental education was especially important in Europe and Israel.

**Interpretation:** This study is the first to apply ML to assess life-course determinants of loneliness during the pandemic across 30 countries. Findings reveal both consistent and context-specific predictors, highlighting the value of early-life information in identifying high-risk groups and guiding targeted, context-sensitive interventions during public health crises.

**Funding:** European Research Council, Academy of Finland, and others.

## 1. Introduction

Loneliness, defined as a subjective negative feeling related to the lack of a wider social network and desired relationships ^1^, is associated with adverse health outcomes, such as chronic inflammation, cardiovascular diseases and early mortality ^2,3^. Studies have shown that loneliness is associated with worse cognition in aging mediated by depression ^4^ and significantly increases the risk of mortality in older adults ^5,6^. With the population rapidly aging worldwide, understanding the consequences and factors behind loneliness in older populations has gained more attention. The COVID-19 pandemic further exacerbated loneliness due to social restriction measures ^7^, emphasizing the need to identify predictors and develop effective interventions for loneliness, especially in times of public health crises.

Previous studies have investigated a wide range of social determinants of loneliness among older adults ^8–11^. However, these studies often have significant limitations. First, most previous research has focused on a narrow set of individual-level social predictors, such as gender ^12^ and education ^13^, while neglecting broader social determinants, such as childhood circumstances, access to healthcare, and economic difficulties. Second, the reliance on traditional statistical models, such as logistic regression ^14–19^, can lead to biased selection of predictors, potentially ignoring complex relationships between social determinants and loneliness. To supplement such approaches, machine learning (ML) methods offer a data-driven approach that can consider a wider range of predictors and address multicollinearity issues ^20^, thus capturing understudied social factors that may influence loneliness. Third, most studies have only looked at current factors, ignoring the life-course perspective that recognizes loneliness as an outcome shaped by experiences at different life stages ^15,21^. While life-course determinants are assessed in some studies ^15,18,22^, only a few studies have utilized ML techniques ^23^, typically using a small number of predictors of loneliness in old age. Finally, previous studies have focused on a particular region, not allowing analyses of how the social determinants of loneliness vary across sociocultural contexts. Although some cross-national comparisons exist ^24,25^, indicating that certain social determinants are consistent across countries and that effective interventions may be transferable, these studies lack a life-course perspective and do not utilize ML techniques.

To address these gaps, we utilized 6 ML models to evaluate 18 life-course social predictors of loneliness across various regional contexts during the COVID-19 pandemic. By including a broad range of social predictors from early life, pre-pandemic, and pandemic periods, we identified the most significant predictors and created a streamlined model to predict loneliness. Our life-course, cross-national approach provides a comprehensive understanding of social determinants of loneliness in late life. This approach may help identify vulnerable segments of the aging population and propose context-sensitive interventions that can be used to alleviate loneliness among aging populations, especially in the event of future public health crises.

## 2. Data and Method

### 2.1. Participants

We used data from three nationally representative surveys: the Health and Retirement Survey (HRS), the English Longitudinal Study of Ageing (ELSA), and Ageing and the Survey of Health, Retirement in Europe (SHARE). Shortly after the outbreak of the COVID-19 pandemic, these surveys started collecting data on the pandemic’s effects on health, known as the HRS COVID-19 survey, ELSA COVID-19, and SHARE COVID-19, respectively. The HRS COVID-19 survey conducted interviews with participants over 50 years old residing in the US in June 2020 or later. The ELSA COVID-19 survey interviewed participants aged 50 and older living in England during the summer of 2020. The SHARE COVID-19 survey interviewed participants aged 50 and older living across 27 European countries and Israel around the summer of 2020. Additionally, we included data from the most recent regular surveys conducted before the onset of the COVID-19 pandemic. More detailed information about these surveys is available from other sources ^26–28^.

We focused on participants aged 65+ and excluded those with missing information on loneliness during the COVID-19 pandemic. The final cohort included 2,851 participants from the HRS COVID-19 survey, 4,478 participants from the ELSA COVID-19 survey, and 39,687 participants from the SHARE COVID-19 survey. Figure 1 illustrates the sample selection process.

**Figure 1.**
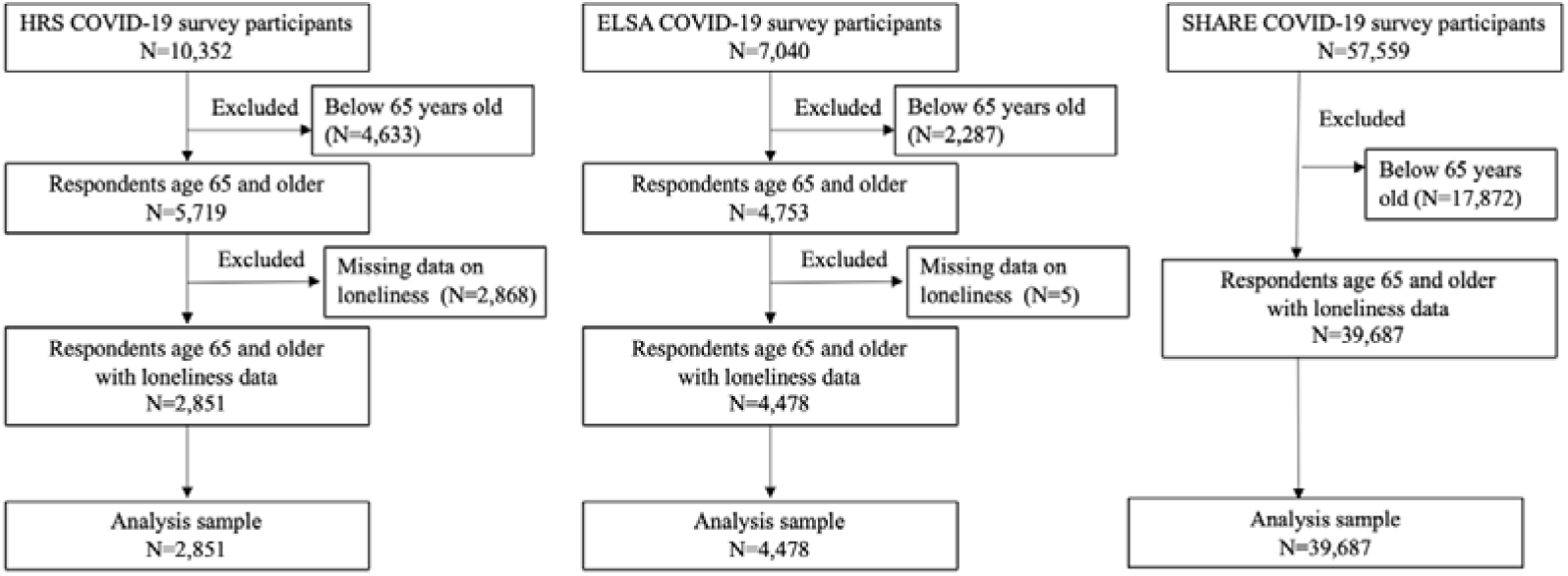
Process of sample selection.

### 2.2. Measures

In the HRS COVID-19 survey, participants were asked to report, “How often do you feel lonely? Often, some of the time, or hardly ever or never?”; in the ELSA COVID-19 survey, participants were asked, “How often do you feel lonely? Often, some of the time, or hardly ever or never”; in the SHARE COVID-19 survey, participants were asked, “How often do you feel lonely? Often, sometimes, or hardly ever or never?”. To conduct a comparative analysis of predicting loneliness among these three surveys, we created a binary variable called loneliness, where “Often, some of the time, or sometimes” was categorized as “feel lonely” while “hardly ever or never” was categorized as “not feel lonely”.

Since a variety of social predictors throughout the life course may impact loneliness during the COVID-19 pandemic, we included 18 predictors from 6 domains: childhood circumstances (father’s education, mother’s education, and occupation type of family breadwinner), demographics (gender, education, marriage, and age), health behaviors (smoking and alcohol consumption), economic situations (receiving pension income before the pandemic, total household income before the pandemic, missing paying bills during the pandemic, and receiving financial support during the pandemic), social connection (weekly contact with children), and COVID-19-related adversity (ever being diagnosed with COVID, anyone died from COVID, delaying medical care, and delaying medical care-surgery). Only predictors with less than 40% missing data were included. Detailed explanations of these predictors can be found in Supplementary material.

### 2.3. Machine Learning Model

Six ML models including CatBoost, XGBoost, Random Forest, Decision Tree, K-Nearest Neighbors (KNN) and Naïve Bayes were utilized to assess the importance of predictors when predicting loneliness. ML models offer several advantages over traditional linear regression models. First, ML excels at capturing non-linear relationships between predictors and outcomes, which linear models often miss. Second, it effectively handles multicollinearity, where predictors are susceptible to being highly correlated. Third, as non-parametric models, ML can identify a wider variety of patterns in the data. All ML analyses were conducted using Python 3.10.9.

#### 2.3.1. Data Pre-Processing

To address missing values of predictors, we used median imputation for continuous variables and mode imputation for categorical variables, which has become a widely used approach for imputing missing values ^29,30^.

#### 2.3.2. Model Development

The process of model development is described in Figure 2. To assess the generalization capability of the ML models, we divided the dataset into training (70%) and testing (30%) subsets. Due to the imbalance in classes, with a minority of participants reporting loneliness (46% in the HRS dataset, 27% in the ELSA dataset, and 31% in the SHARE dataset), we adjusted the scale_pos_weight parameter of the ML model accordingly. Hyperparameter tuning was conducted using both random search and grid search methods with 10-fold cross-validation to identify optimal hyperparameters for ML models, an approach shown in prior studies to improve ML performance and generalizability ^31,32^.

**Figure 2.**
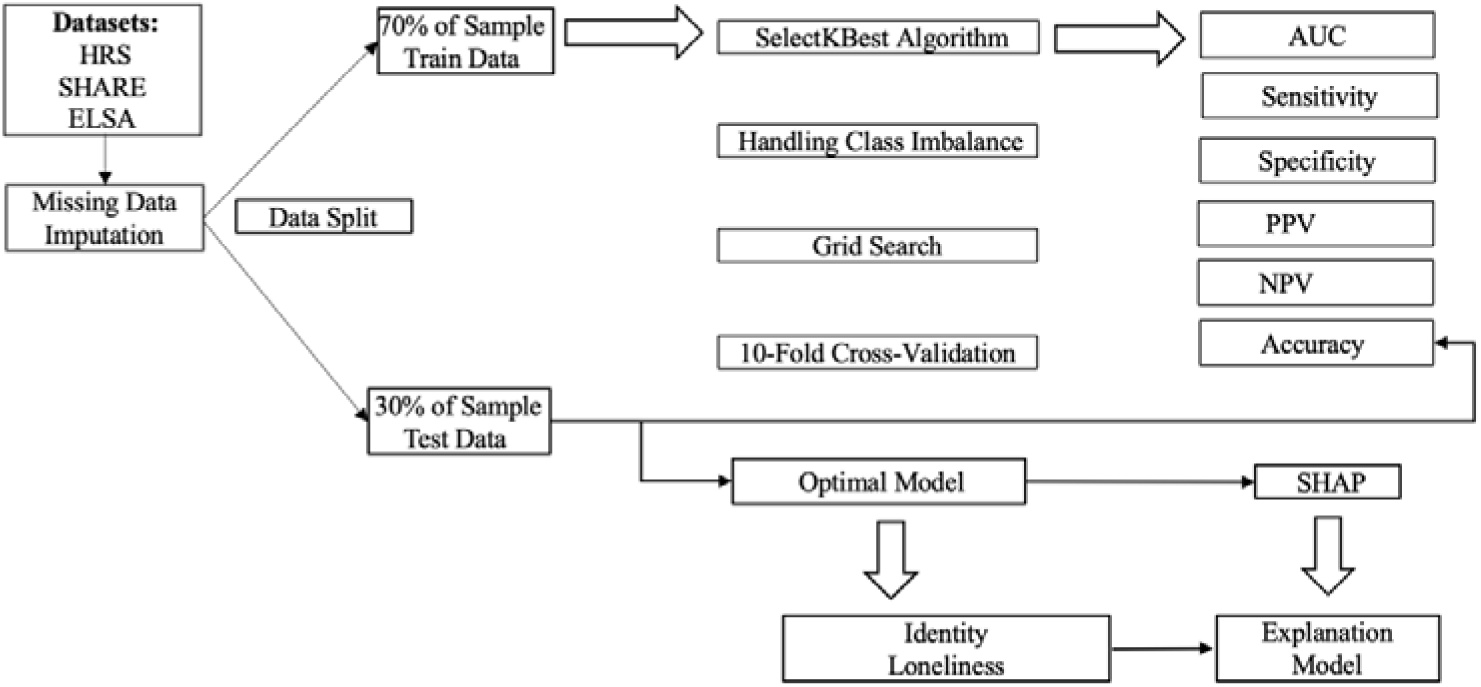
Process of model development.

The lack of interpretability is a significant shortcoming of ML models. Recently, Shapley Additive Explanations (SHAP), an extension of game theory, have been utilized in studies of ML-based predictions to measure the contribution of each predictor in these models^33^. In this study, SHAP values is used to quantify both the magnitude and direction of the associations between predictors and loneliness. Moreover, the distribution and clustering of SHAP values along the x-axis may indicate potential non-linear relationships between each predictor and loneliness.

The predictive performance of the ML models was evaluated using a set of metrics, including the area under the curve (AUC), accuracy, positive predictive value (PPV), negative predictive value (NPV), sensitivity, and specificity.

## 3. Result

### 3.1. Descriptive Analysis

Characteristics of participants in the HRS (n=2,851, 60% women), ELSA (n=4,478, 55% women), and SHARE (n=39,687, 57% women) are presented in Table 1. The mean (standard deviation (SD)) age of participants was 75.40 (7.50) years in the HRS, 73.96 (6.55) years in ELSA, and 74.63 (7.04) years in SHARE. During the COVID-19 pandemic, the prevalence of loneliness varied across regions, with 46% of participants reporting loneliness in the US (HRS dataset), 27% in England (ELSA dataset), and 31% in Europe and Israel (SHARE dataset).

**Table 1.** Demographic characteristics and loneliness of study participants.

|  | HRS | ELSA | SHARE |
| --- | --- | --- | --- |
| Loneliness<br>(Ref. = No) | 1,317 (46%) | 1,295 (27%) | 12,187 (31%) |
| Gender<br>(Ref. = Man) | 1,703 (60%) | 2,603 (55%) | 22,445 (57%) |
| Being married or partnered<br>(Ref. = No) | 1,685 (59%) | 3,362 (71%) | 23,714 (60%) |
| Age (SD) | 75.40 (7.50) | 73.96 (6.55) | 74.63 (7.04) |

Note: Values are presented as n (%) unless otherwise indicated. n indicates the sample size and % indicates the proportion within each dataset. Age is shown as mean (standard deviation, SD). “Ref.” indicates the reference category.

### 3.2. Model Performance

CatBoost demonstrated the best predictive ability on the test data for predicting loneliness among older adults during the COVID-19 pandemic across all datasets (Figure 3). In the HRS dataset, XGBoost achieved an accuracy of 0.632, AUC of 0.603, sensitivity of 0.581, specificity of 0.620, PPV of 0.558, and NPV of 0.642. In the ELSA dataset, the accuracy was 0.693, AUC 0.641, sensitivity 0.641, specificity 0.641, PPV 0.406, and NPV 0.823. In the SHARE dataset, CatBoost achieved the highest accuracy of 0.718, with an AUC of 0.672, sensitivity of 0.657, specificity of 0.678, PPV of 0.477, and NPV of 0.816. These findings suggest that loneliness prediction models perform better in Europe and Israel than in England and the US.

**Figure 3.**
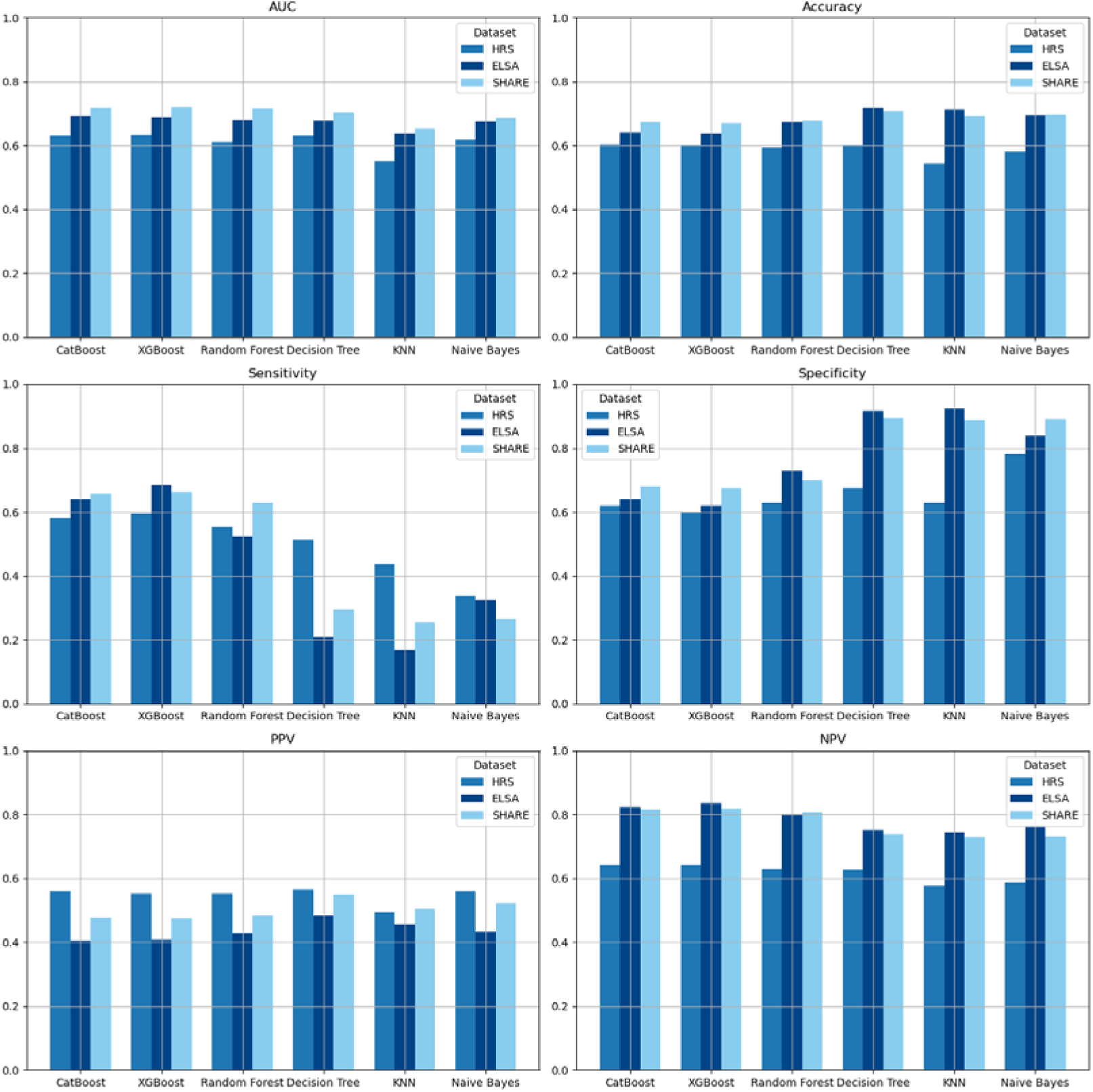
Predictive performance of ML models in this study.

### 3.3. Predictor Importance

#### Common Predictors across Datasets

Several social predictors consistently ranked as the top predictors across all datasets (Figure 4), underscoring their robustness in predicting loneliness during the COVID-19 pandemic. Marital status emerged as the most influential predictor across all three datasets, highlighting that being married or partnered contributed to reducing the likelihood of experiencing loneliness. Women were more prone to loneliness. Childhood circumstances, including parental education and breadwinner occupation, also emerged as strong predictors of loneliness. For instance, a lower-status breadwinner occupation during childhood was associated with a higher risk of loneliness in both England and Europe and Israel. COVID-19-related adversities, such as delayed medical care during the pandemic, were closely linked to an increased likelihood of loneliness in the US, as well as in Europe and Israel.

**Figure 4.**
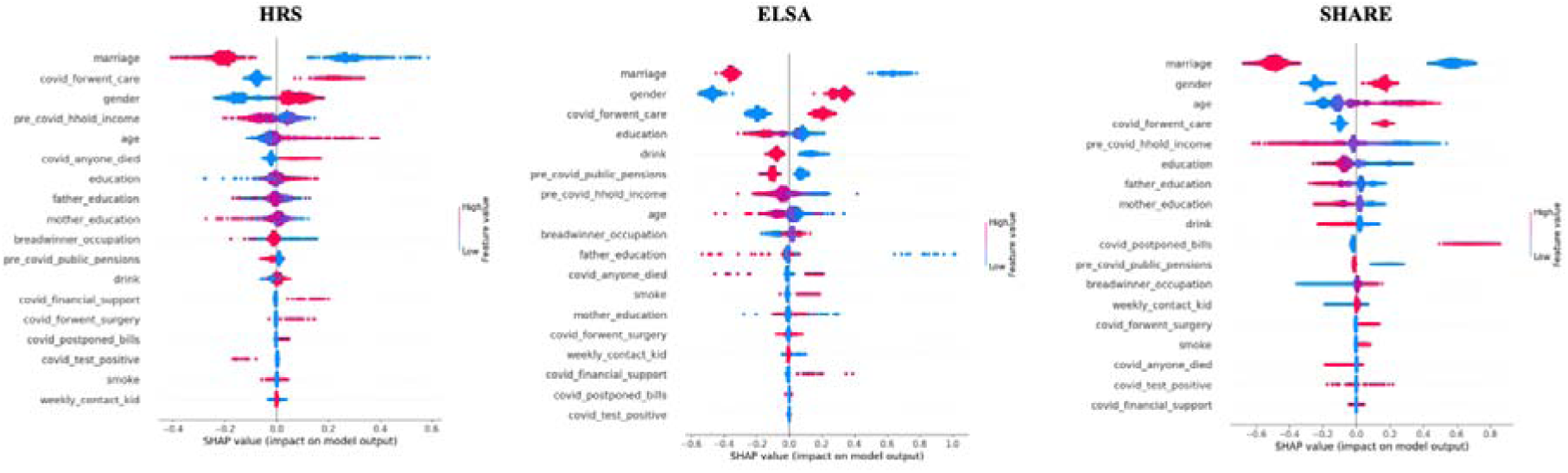
Importance of life-course social predictors for loneliness Note: SHAP values indicate the magnitude and direction of these associations, with blue points meaning lower values and red points meaning higher values for the conditions on the horizontal axis. They show the importance ranking on the vertical axis, with higher values meaning higher importance. Additionally, the distribution and clustering of SHAP values along the x-axis means potential non-linear relationships between each predictor and the outcome, indicating that the impact on the outcome is not constant.

#### Distinct Predictors across Regions

In the US (HRS dataset), delaying medical care during the pandemic was identified as a more crucial predictor than in England, Europe, and Israel, highlighting the role of healthcare insecurity in loneliness among older Americans. Additionally, economic factors, such as pre-pandemic household income and receiving financial support from others during the pandemic, were found to be important predictors of loneliness, suggesting the role of financial constraints in loneliness among older adults in the US.

In England (ELSA dataset), education was found to be a stronger predictor than in the US, Europe and Israel, with higher levels of education generally associated with a lower odds of loneliness, highlighting its protective role. Additionally, receiving a public pension before the pandemic and the occupation type of family breadwinner during childhood were identified as more significant predictors.

In Europe and Israel (SHARE dataset), age was identified as a more significant predictor, indicating that older adults are more likely to experience feelings of loneliness. Additionally, parental education emerged as a more important social predictor, with higher education indicating a lower risk of loneliness. These findings suggest the lasting influence of childhood circumstances on late-life loneliness among European and Israeli older adults.

### 3.4. Non-linear Relationships Between Predictors and Loneliness

SHAP dependency plots revealed several non-linear associations between social determinants and loneliness (Figure 5). In all three datasets, there were non-linear associations between age and loneliness among older adults. For example, in the SHARE dataset, loneliness risk sharply increased after the age of around 80. Education levels also showed a non-linear relationship with late-life loneliness. For instance, in the ELSA dataset, completing education at the age of 18 and above (education value of 5 and 6 in Figure 4) was associated with lower loneliness risk. In the SHARE dataset, a threshold effect was observed at around 13 or more years of education, beyond which higher education levels were linked to lower loneliness risk.

**Figure 5.**
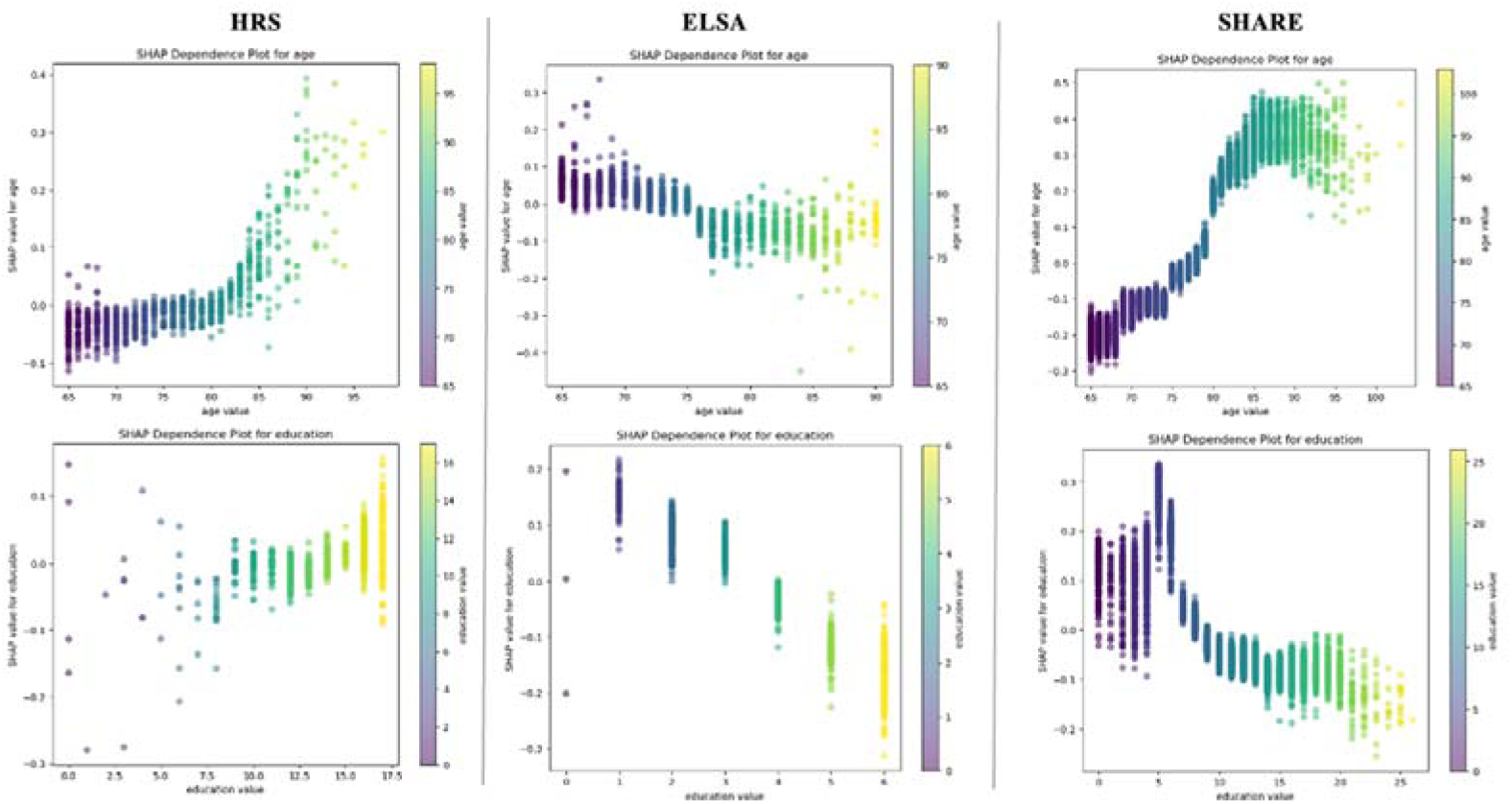
SHAP dependency plots for age or education levels and loneliness Note: The x-axis represents different levels of age or education, while the y-axis indicates the SHAP values, which quantify the contribution of each age or education level to loneliness. Higher SHAP values mean a stronger positive impact on loneliness.

### 3.4. Streamlined Predictive Models

We used a feature selection method to identify the top 10 social predictors for constructing streamlined predictive models across regions (Figure 6). These models achieved predictive performance close to that of the full 18-predictor model. This enables us to streamline the prediction model, effectively assess the risks of loneliness and implement interventions to reduce loneliness among older adults in future public health crises. The top 10 predictors varied by region: in the US, they included marital status, pre-pandemic household income, age and other predictors; in England, marital status, gender, delayed medical care during the pandemic, and other predictors; and in Europe and Israel, marital status, pre-pandemic household income, age, and other predictors. The complete list of the top 10 predictors is provided in the note for Figure 6.

**Figure 6.**
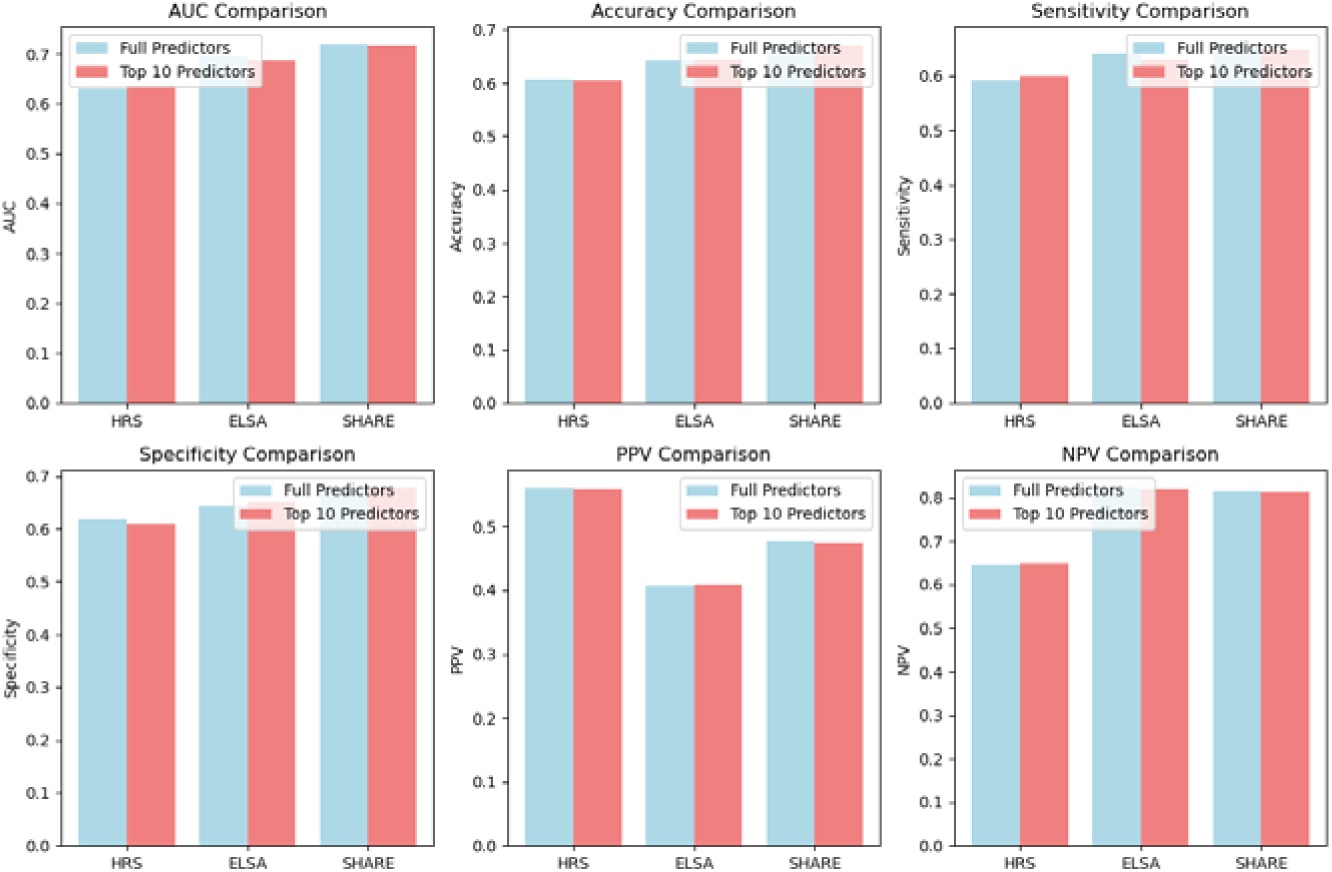
Predictive performance of ML models using full predictors and top 10 predictors Note: In the HRS dataset, the top 10 predictors include: marital status, pre-pandemic household income, age, mother education levels, education levels, delaying medical care during the pandemic, gender, father education levels, breadwinner occupation during childhood, anyone died from COVID-19; in the ELSA dataset, the top 10 predictors include: marital status, gender, delaying medical care during the pandemic, receiving public pensions before the pandemic, household income before the pandemic, education levels, drinking alcohol, breadwinner occupation during childhood, age, father education levels; in the SHARE dataset, the top 10 predictors include: marital status, pre-pandemic household income, age, gender, education levels, delaying medical care during the pandemic, postponing bills during the pandemic, mother education levels, father education levels, drinking alcohol.

## 4. Discussion

To the best of our knowledge, this is the first cross-national study on the life-course predictors of loneliness during the COVID-19 health crisis. By incorporating a life-course perspective and a comprehensive set of social predictors spanning from early-life, pre-pandemic, and pandemic periods, our findings enhance our understanding of how social determinants shape loneliness across various sociocultural contexts during the global health crisis.

### Key Social Predictors of Loneliness

Our results indicate that certain social determinants were consistent across regions, while others varied depending on the national contexts. Marital status was the strongest predictor in the US, England, Europe and Israel. Older adults who were married or partnered had more social support ^34^, and hence were less likely to experience loneliness, emphasizing the protective role of marriage against loneliness in both pandemic and non-pandemic settings ^35–37^. Gender was another critical predictor, with women showing a higher likelihood of loneliness across all regions, likely because they tend to be widowed and live alone ^38^, consistent with previous studies highlighting gender differences in loneliness ^39,40^. Childhood circumstances, particularly parental education and breadwinner occupation, also played an important role in predicting loneliness. This underscores the lasting impact of early-life socioeconomic status on late-life loneliness ^41–43^. Similarly, COVID-19-related adversity, such as delaying medical care during the pandemic, was a consistent predictor in all regions, indicating the immediate impact of the pandemic on loneliness.

While these social predictors were common across regions, the relative importance of several of them varied. In the US, the economic situation, such as pre-pandemic household income and receiving financial support from others during the pandemic, was a more important predictor. This highlights the financial constraints of older Americans and their influence on loneliness ^44^. In England, receiving a public pension before the pandemic and the occupation type of the family breadwinner during childhood were more crucial, suggesting the long-term economic influences on loneliness in this context. In Europe and Israel, parental education ranked as a stronger predictor, emphasizing the enduring impact of parental education disparities on children’s late-life loneliness.

We further explored the non-linear relationships between age, education levels, and loneliness, highlighting the complexity of these connections. Specifically, we discovered threshold effects where the risk of loneliness significantly increased after the age of around 80 in Europe and Israel, highlighting the heightened vulnerability of the oldest-old to loneliness. These findings are consistent with prior research that indicates a non-linear relationship between age and loneliness in non-pandemic settings ^45^. Similarly, the relationship between education and loneliness was also non-linear, with protective effects observed beyond certain education thresholds in England, Europe, and Israel.

### Model Predictive Performance

Our findings demonstrate that the ML methods, particularly the CatBoost model, effectively predicted loneliness with varying levels of accuracy across different regions. The model achieved the highest AUC among older adults in Europe and Israel, followed by England and the US. These variations may reflect differences in sociocultural influences. The relatively lower performance in England and the US may be due to the absence of additional unmeasured key predictors in our models that could influence loneliness in these countries, such as race, ethnicity, and neighborhood social cohesion, all of which have been shown to be associated with loneliness among older adults during the pandemic in the US and England ^46–48^.

### Streamlined Predictive Model for Future Applications

We developed a streamlined predictive model using the top 10 most influential social predictors. This streamlined model maintains high predictive accuracy while reducing the complexity of variable selection, making it a practical tool for future public health crises. Interestingly, although COVID-19-related adversity influenced loneliness, only a few of those items were among the top 10 predictors. This suggests that loneliness during the COVID-19 pandemic was primarily influenced by demographics (e.g., age, marital status, and education) rather than the immediate effects of the crisis. While prior studies have largely examined demographics and pandemic-related factors separately ^41,49^, our study integrates both perspectives and demonstrates that demographics played a more dominant role in shaping loneliness during COVID-19.

### Strengths and Limitations

This study has strengths in four aspects. First, it utilized data from large, nationally representative samples across 28 European countries, the US, and Israel, allowing for robust cross-national comparisons. The harmonized structure of the HRS, ELSA, and SHARE facilitated direct comparisons of loneliness predictors across different sociocultural contexts. Second, it used a comprehensive set of life-course social predictors and identified their contributions to loneliness, not only including well-established factors identified in prior research but also exploring lesser-studied factors such as childhood circumstances that may contribute to later-life loneliness. This can provide insights for creating targeted measures to reduce the loneliness of vulnerable older adults. Third, the application of ML models overcame the limitations of traditional statistical models widely used in previous studies and hence provided us with deeper understanding of dealing with loneliness. Fourth, we developed a streamlined predictive model that maintains similar accuracy while reducing the number of predictors. This streamlined approach advances the model’s usability for practical applications in future public health crises.

However, there are still some limitations in this study. First, sociocultural factors affecting how loneliness is reported may influence differences across regions. Second, our study focuses on predictive modeling rather than causal inference, limiting our ability to establish direct causal relationships between life-course social determinants and loneliness. Third, our datasets are restricted to the early stage of the pandemic (April to August 2020), which may not fully capture potential heterogeneity across different phases of the pandemic. Additionally, our datasets used in this study are from high-income countries, restricting the generalizability of our findings to lower-income settings. Finally, although we incorporated a wide range of life-course predictors, data availability and the need for cross-national comparability meant that some important factors remained unmeasured—for example, race, which may be particularly relevant in the US and could reduce the predictive performance of the ML model in this context ^47^.

## 5. Conclusion

In conclusion, our findings indicate the importance of life-course social determinants in shaping loneliness in late life and emphasize the significance of context-sensitive interventions. While some early-life factors are not directly modifiable, they can serve as important indicators for identifying individuals who may be particularly vulnerable to loneliness. By identifying key predictors across various national contexts, this study highlights the need for tailored prevention strategies and broader efforts to identify and support high-risk segments of the population to address loneliness, especially in anticipation of future public health crises. Future research should explore the causal pathways underlying these associations and expand this research to diverse global contexts to ensure that loneliness prevention measures are effective and equitable.

## Supporting information

Detailed description of social predictors of loneliness

## Data Availability

The datasets analyzed in this study, including HRS, SHARE, and ELSA, are publicly available from the following websites: https://hrs.isr.umich.edu/, https://share-eric.eu/, and https://www.elsa-project.ac.uk/, respectively.

https://share-eric.eu/

https://www.elsa-project.ac.uk/

