## Supplementary material for "Machine Learning to Investigate Life-Course Social Determinants of Loneliness among Older Adults in the US, England, Israel, and 27 European Countries during the Pandemic": Detailed description of social predictors of loneliness: 2.2. Supplementary material.docx

Detailed description of social predictors of loneliness

| **Domain** | **Social Predictors** | **Meaning of Social Predictors** | | |
| --- | --- | --- | --- | --- |
|  |  | **HRS** | **ELSA** | **SHARE** |
| Demographics | gender | Participant’s gender:   1. Man 2. Woman | Participant’s gender:  1. Man  2. Woman | Participant’s gender:  1. Man  2. Woman |
|  | education | Years of education attained by the participant: ranging from 0 to 17 years | Age at which the participant completed their education:   0. None 1. Aged 14 or under 2. Age 15 3. Age 16 4. Age 17 5. Age 18 6. Age 19 or over | Years of education attained by the participant: ranging from 0 to 38 years |
|  | marriage | Participant’s marital status:   0. Separated; divorced, widowed; never married 1. Married; married, spouse absent; partnered | Participant’s marital status:   0. Separated; divorced, widowed; never married 1. Married; partnered | Whether the participant is married or partnered:   0. No 1. Yes |
|  | age | Participant’s age: 65 years or older | Participant’s age: 65 years or older | Participant’s age: 65 years or older |
| Health Behaviors | smoke | Whether the participant smokes now:   0. No 1. Yes | Whether the participant smokes now:   0. No 1. Yes | Whether the participant smokes now:   0. No 1. Yes |
|  | drink | Whether the participant drinks any alcohol:   0. No 1. Yes | Whether the participant drinks any alcohol every week:   0. No 1. Yes | Whether the participant drank alcohol in the last 7 days:   0. No 1. Yes |
| Economic Situations | pre_covid_public_pensions | Whether the participant was currently receiving pension income before the COVID-19 pandemic:   0. No 1. Yes | Whether the participant was currently receiving pension income before the COVID-19 pandemic:   0. No 1. Yes | Whether the participant received public pension before the COVID-19 pandemic:   0. No 1. Yes |
|  | pre_covid_hhold_income | Total household income of the participant before the COVID-19 pandemic ranges from 0 to 1,324,240 and is log-transformed for analysis. | Total couple income of the participant before the COVID-19 pandemic ranges from 0 to 466,100.9688 and is log-transformed for analysis. | Total household income of the participant before the COVID-19 pandemic ranges from 0 to 1.24e+07 and is log-transformed for analysis. |
|  | covid_postponed_bills | Whether the participant missed paying their bills during the COVID-19 pandemic:   0. No 1. Yes | Whether the participant took a bill payment holiday during the COVID-19 pandemic:   0. No 1. Yes | Whether the participant postponed the bills during the COVID-19 pandemic:   0. No 1. Yes |
|  | covid_financial_support | Whether the participant received financial support from others during the COVID-19 pandemic:   0. No 1. Yes | Whether the participant received financial support from others during the COVID-19 pandemic:   0. No 1. Yes | Whether the participant received financial support from others during the COVID-19 pandemic:   0. No 1. Yes |
| Social Connection | weekly_contact_kid | Whether the participant has any weekly contact with their children in person/phone/mail/email:   0. No 1. Yes | Whether the participant has any weekly contact with their children or relative by phone/video:   0. No 1. Yes | Whether the participant has any weekly contact with their children by phone/e-mails:   0. No 1. Yes |
| COVID-19-related Adversity | covid_test_positive | Whether the participant was ever diagnosed with COVID:   0. No 1. Yes | Whether the participant tested positive for coronavirus:   0. No 1. Yes | Whether the participant tested positive for COVID-19:   0. No 1. Yes |
|  | covid_anyone_died | Whether anyone close to you died from COVID:   0. No 1. Yes | Whether anyone close to you died from COVID:   0. No 1. Yes | Whether anyone close to you died from COVID:   0. No 1. Yes |
|  | covid_forwent_care | Whether the participant delayed medical care:   0. No 1. Yes | Whether the participant was unable to access needed medical or dental care:   0. No 1. Yes | Whether the participant forwent/postponed/denied medical care:   0. No 1. Yes |
|  | covid_forwent_surgery | Whether the participant delayed medical care-surgery:   0. No 1. Yes | Whether the participant had an operation or treatment cancelled:   0. No 1. Yes | Whether the participant forwent/postponed/denied medical care-treatment:   0. No 1. Yes |
| Childhood Circumstances | father_education | Years of education attained by participant’s father: ranging from 0 to 17 years | Age at which the participant’s father completed his education:   1. Never went to school 2. Aged 14 or under 3. Age 15 4. Age 16 5. Age 17 6. Age 18 7. Age 19 or over | Father’s education by isced code:   0. None 1. Primary education 2. Lower secondary education 3. Upper secondary education 4. Post-secondary non-tertiary education 5. First stage of tertiary education 6. Second stage of tertiary education |
|  | mother_education | Years of education attained by participant’s mother: ranging from 0 to 17 years | Age at which the participant’s mother completed her education:   1. Never went to school 2. Aged 14 or under 3. Age 15 4. Age 16 5. Age 17 6. Age 18 7. Age 19 or over | Mother’s education by International Standard Classification of Education (ISCED) code:   0. None 1. Primary education 2. Lower secondary education 3. Upper secondary education 4. Post-secondary non-tertiary education 5. First stage of tertiary education 6. Second stage of tertiary education |
|  | breadwinner_occupation | Father’s occupation when the participant was 16 years old:  1. White-collar  2. Blue-collar  3. Military | Main carer’s occupation when the participant was 14 years old:  1. White-collar  2. Blue-collar  3. Military  4. Other | Main breadwinner’s occupation when the participant was 10 years old:  1. Legislator, senior official or manager  2. Professional  3. Technician or associate professional  4. Clerk  5. Service, shop or market sales worker  6. Skilled agricultural or fishery worker  7. Craft or related trades worker  8. Plant/machine operator or assembler  9. Elementary occupation  10. Armed forces  11. Spontaneous only: there was no main breadwinner |
